# Fast spread of COVID-19 in Europe and the US suggests the necessity of early, strong and comprehensive interventions

**DOI:** 10.1101/2020.04.04.20050427

**Authors:** Ruian Ke, Steven Sanche, Ethan Romero-Severson, Nick Hengartner

## Abstract

The COVID-19 pandemic caused more than 800,000 infections and 40,000 deaths by the end of March 2020. However, some of the basic epidemiological parameters, such as the exponential epidemic growth rate and *R*_0_ are debated. We developed an inference approach to control for confounding factors in data collection, such as underreporting and changes in surveillance intensities, and fitted a mathematical model to infection and death count data collected from eight European countries and the US. In all countries, the early epidemic grew exponentially at rates between 0.19-0.29/day (epidemic doubling times between 2.4-3.7 days). This suggests a highly infectious virus with an *R*_0_ likely between 4.0 and 7.1. We show that similar levels of intervention efforts are needed, no matter the goal is mitigation or containment. Early, strong and comprehensive intervention efforts to achieve greater than 74-86% reduction in transmission are necessary.

**One-sentence Summary:** We estimated that COVID-19 spreads rapidly in 8 European countries and the US, suggesting that early, strong and comprehensive interventions are necessary.

## Introduction

COVID-19 originated in Wuhan China in Dec, 2019 (*1*). It has spread rapidly and caused a global pandemic within a short period of time. As of March 31, 2020, the global pandemic lead to more than 800,000 total confirmed cases and 40,000 deaths. Estimation of the rate of early epidemic spread in Wuhan, China, lead to different conclusions. Initially, it was suggested that the epidemic grew at 0.1-0.14/day, leading to an epidemic doubling time of 5-7 days (*2-5*). However, using domestic travel data and two distinct approaches, we estimated that the epidemic in Wuhan grew much faster than initially estimated, and the growth rate is likely to be between 0.21-0.3/day, translating to a doubling time between 2.3 to 3.3 days, and an *R*_0_ approximately at 5.7 with a large confidence interval (*6*). A fast epidemic spread is consistent with multiple other lines of evidence, such as the rapid increase of the epidemic curve by symptom onset published by China CDC (*7*) and the growth in the number of death cases in Hubei, China during late January 2020 (*6*). However, it was not clear whether COVID-19 can spread in other countries as fast as in Wuhan, China.

Accurate estimation of the rate of epidemic growth is important for many practical aspects. First, it is crucial for forecasting the epidemic trajectory, the burden on health care systems and potential health and economic damage, so that appropriate and timely responses can be prepared. Second, it sets the baseline for evaluation of effectiveness of intervention strategies. Third, it is important for accurate estimation of the basic reproductive number, *R*_0_, which in turn is used for many control measures, including evaluation of the vaccine/herd immunity threshold needed to stop transmission (*6, 8*). However, a major challenge to the inference of the growth of COVID-19 is that as a result of a fast-growing outbreak and a sizable infected population with no or mild-to-moderate symptoms (*9, 10*), case confirmation data is influenced by many factors in addition to the true epidemic growth, including substantial underreporting (*11*), i.e. low detection rate, changes in surveillance intensity and delays in case confirmation. Simply fitting an exponential curve to case confirmation data may lead to erroneous conclusions when confounding factors are not taken into account or estimated from other sources of data.

Here, we argue that death and the cause of death are usually recorded reliably and are less affected by surveillance intensity changes or delay in confirmation than case counts. The time series of death counts reflects the growth of an epidemic reliably, with a delay in onset determined by the time between infection to death. Although it is possible that deaths from COVID-19 were not recorded as death from COVID-19 during very early phase of an epidemic when people are unaware of community transmission of COVID-19 (*12*) or during the relatively late phase of the epidemic when health care system is overwhelmed, this represents only a small fraction of cases in the countries we examine in this work, and is unlikely to significantly affect the death count curve when the death count increases exponentially. Here, we designed a simple methodology to disentangle the epidemic growth from confounding factors, such as underreporting, delays in case confirmation and changes in surveillance intensity. We fit models to both case incidence data and death count data collected from eight European countries and the US in March 2020. We show that in most countries, the detection rate of infected individuals is in general low, and COVID-19 spreads very fast in these countries. For such a fast-epidemic growth, our results suggest that very strong and active control measures need to be implemented as early as possible regardless of the public health goal (e.g. mitigation versus containment), and that moderate control measures will not achieve measurable public health benefit.

## Methods

### Data

We collected daily case confirmation and death count data for the US and 8 countries in Europe from the John Hopkins CSSE (Center for Systems Science and Engineering) database (https://github.com/CSSEGISandData/COVID-19). The data is accessed and extracted on March 31, 2020. The data consists of time series of numbers of case confirmations and deaths by country (cumulative). Daily incidences were derived from the cumulative counts. We used data from the following countries: France (FR), Italy (IT), Spain (SP), Germany (GR), Belgium (BE), Switzerland (SW), Netherlands (NT), United Kingdom (UK) and the US (US).

We included a subset of case confirmation and death count data for inference based on the two following criteria. First, to minimize the impact of stochasticity and uncertainty in early data collection, we used case confirmation incidence data starting from the date when the cumulative number of cases was greater than 100, and used daily new death count data starting from the date when the cumulative death count is greater than 20 in each country.

Second, to estimate the early outbreak growth in each country before control measures were implemented and at the same time maximize the power of inference, we allowed a maximum of 15 days of data points for the two types of data, leading to a maximum of 30 data points for each country. Note that the end date of incidence data used for inference is at or close to the date when strong control measures were implemented in each country (Table S1). We tested the sensitivity of model predictions when only 10, 13 days of data points are included for inference. The results are robust to this variation (Table S1), suggesting that the choice of 15 days is reasonable and that the data shows consistent exponential growth during this period.

### Model

We construct a SEIR type model using ordinary differential equations (ODEs; see Supplementary materials). We consider the exponentially growing phase of the outbreak and thus make the common assumption that the susceptible population is constant over time. Then, the total number of infected individuals *I\** (*t*) *= E*(*t*) + *I*(*t*) can be expressed as:

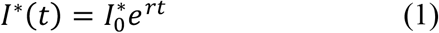

where *r* is the exponential growth rate of the epidemic (the growth rate for short below), and 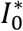 is the number of total infected individual at time 0, set arbituarily as January 20, 2020. Note the choice of the date of time 0 does not affect our estimation.

We solve the ODE model and derive the following expressions for the key quantities for model inference (see Supplementary material). The descriptions and values used for the parameters in the ODE model are summarized in Table 1.

**Table 1.**
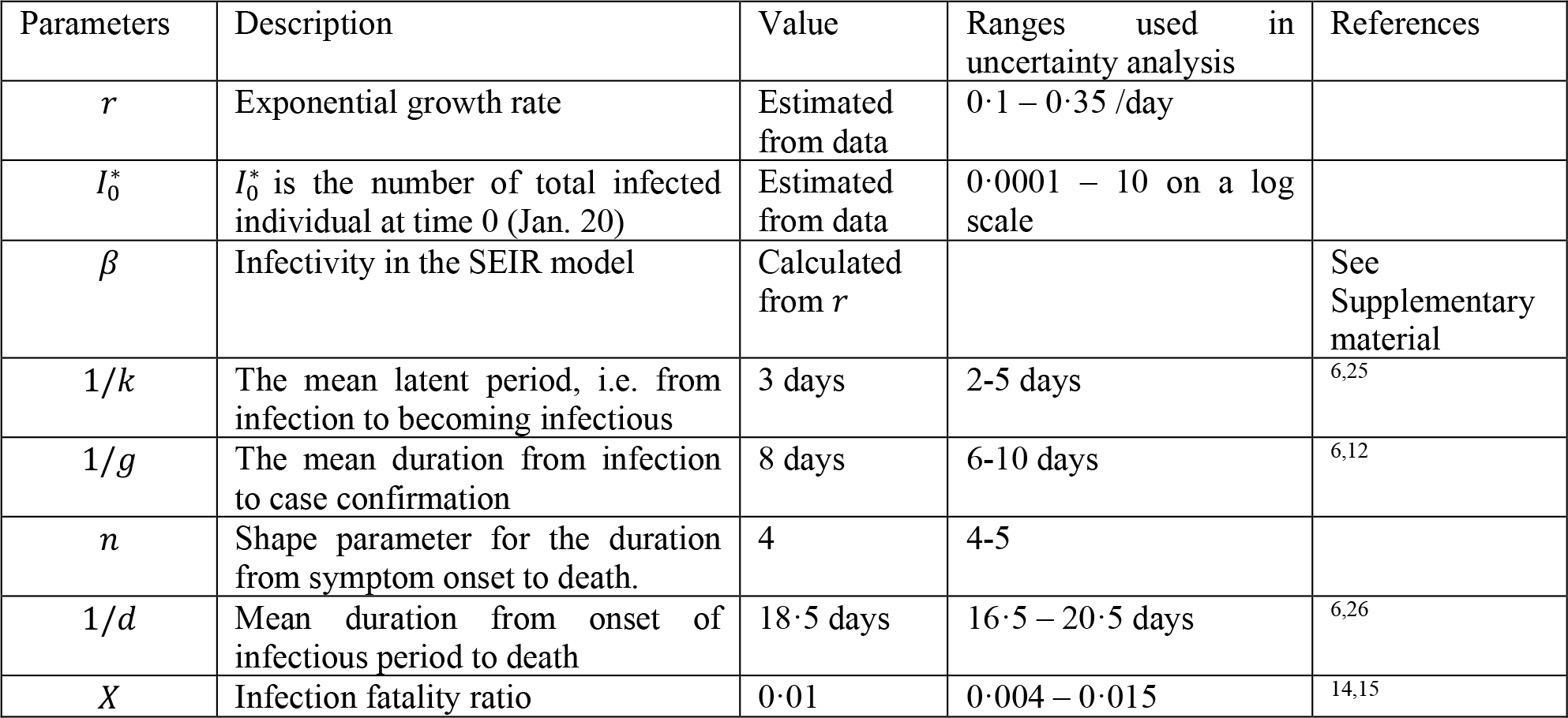
Description of parameter and their values. See the supplementary material for discussions of choice of parameter values.

The true daily incidence of infected individuals, Ω(*t*), is:

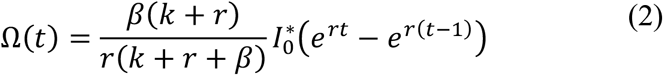

where *β* and 1/*k* are the transmission potential of the virus and the latent period of infection, respectively.

The daily new confirmed case count, Ψ(*t*), is related to the true daily incidence, Ω(*t*) as:

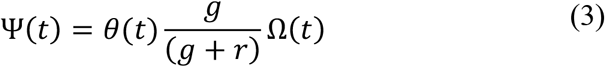

where *θ*(*t*) is the detection rate, i.e. the fraction of newly individuals at time *t* who are later detected by surveillance later on. We assumed a realistic (Erlang) distribution for the period between infection and case confirmation(*6*), where 1/g and *m* are the mean and the shape parameter for the distribution.

The daily new death count, *Φ*(*t*), is related to the true daily incidence, Ω(*t*) as:

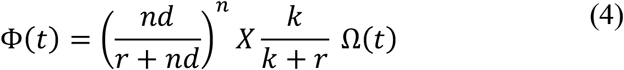

where *X* is the infection fatality ratio (sometimes referred as case fatality ratio, CFR, depending on definition). Again, we assumed a realistic (Erlang) distribution for the period between infection and death (*6*), where 1/*d* and *n* are the mean and the shape parameter for the distribution.

The expressions above clearly establish that during the exponential growth of an epidemic, the ratio between death counts *Φ* and the confirmed cases Ψ (i.e. two widely reported numbers in public databases, publications and news reports) is not only dependent on infection fatality ratio and the detection rate, but also a highly nonlinear relationship of the growth rate, *r* with the distributions of the delay in case confirmation and the period from infection to death. Failure to take this into account may lead erroneous conclusions.

We tested three different scenarios for surveillance intensity changes over time, modeled as the detection rate, *θ*(*t*):

1. *θ* is a constant, i.e. no change over time;
2. 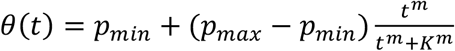 i.e. *θ* is a Hill-type function of *t*;
3. *θ*(*t*) is equal to *p*_min_ before *t*_1_, increases linearly to *p*_max,-_ between *t*_1_ and *t*_2_ and stay constant at *p*_max,-_ after *t*_2_, i.e. *θ* is a semi-linear function of *t*.

Note that the time from infection to case confirmation, 1/g, can be a time dependent function as we and others shown previously(*6, 13*). To keep the model simple, we implicitly assume that the time dependent changes in *g* can be included in the estimation of *θ*(*t*).

See supplementary materials for details of data collection, modeling analysis, parameter choice and estimation and uncertainty quantifications.

## Results

### Estimation of the epidemic growth rate and surveillance intensity

We constructed an SEIR type model and fitted the model to both the incidence (case confirmation) data and the daily new death count data from eight countries in Europe and the US. We selected data from a period during early outbreak before or a few days after strong control measures, such as school and work closure, and locking down cities etc., were implemented in these countries (see Methods for details and Table S1). There are clear decreases in the rate of exponential growth of case incidence after the end dates of data selection in most of these countries (Fig. 1), and this is likely to reflect the impact of the strong control measures implemented (*14*).

**Figure 1.**
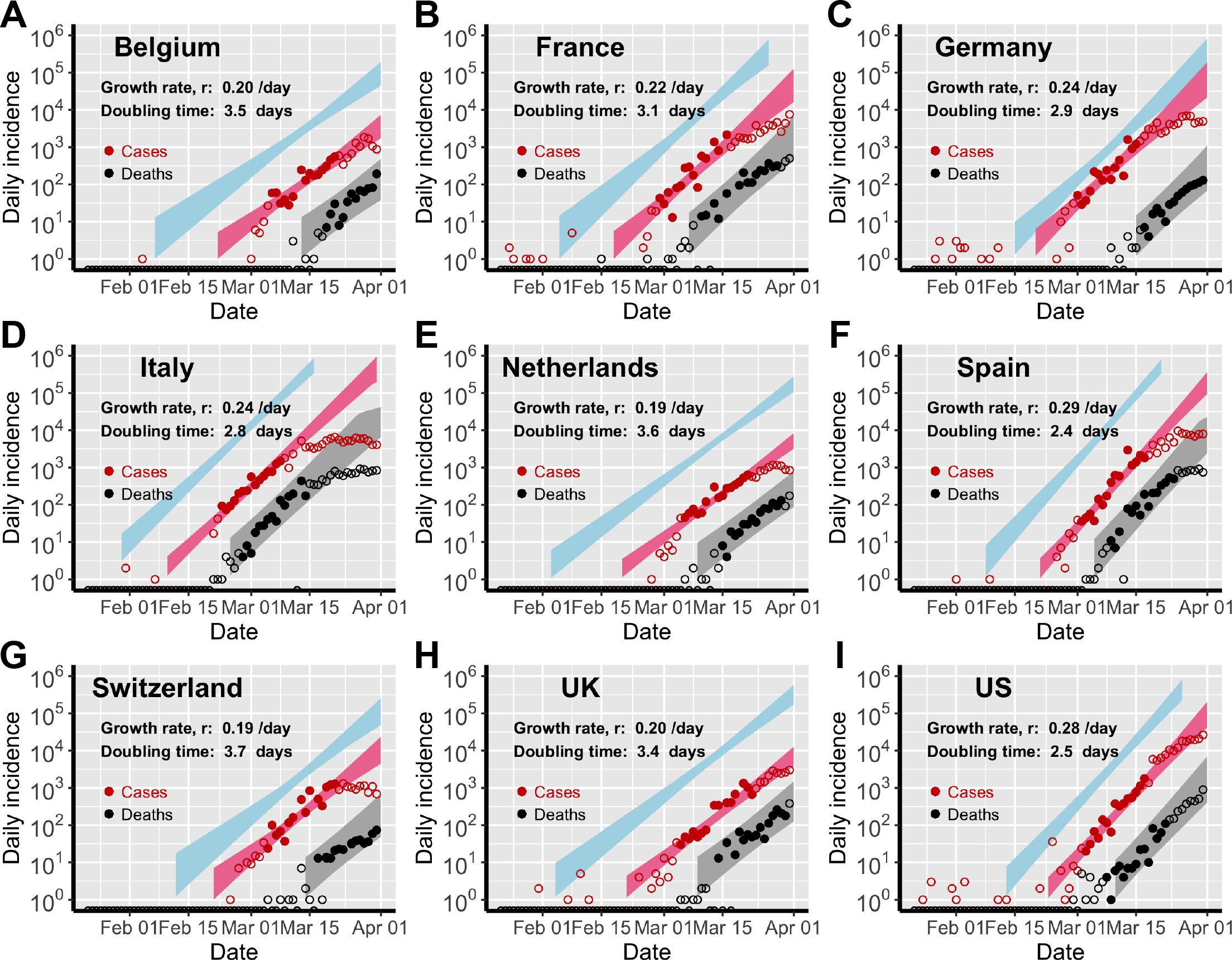
Estimation of the exponential growth rate, doubling time of epidemics and detection rate of infected individuals in 8 European countries and the US. Red and black symbols show the daily counts of new case confirmation and new death, respectively. Dots denote data used for parameter inference; whereas open circles denote data that are not used for parameter inference. We simulated the exponential model using sampled parameter combinations that are able to explain the data shown in dots (see the Uncertainty Quantification section in the Supplementary Material). The colored bands denote the area between the lower and upper bounds of simulated/predicted true daily infection incidence (blue), daily case confirmation (red) and daily death (grey) assuming no intervention efforts nor changes in surveillance intensity. Deviation of open circles from the corresponding bands thus indicates either changes in surveillance intensity or impacts of control measures.

We estimated that the exponential growth rate of early outbreaks, *r*, ranges between 0.19 and 0.29/day in the nine countries, translating to doubling times between 2.4-3.7 days (Fig. 1). The two countries with the highest point estimates of the exponential rate are Spain and the US, at 0.29 and 0.28/day, respectively; whereas Switzerland and Netherlands have lower point estimates at 0.19/day. Accounting for uncertainties in the estimated parameter values (see Methods), we found that the epidemic growth rates in France, Germany, Italy, Spain and the US are mostly likely higher than 0.2/day (Fig. 2A).

**Figure 2.**
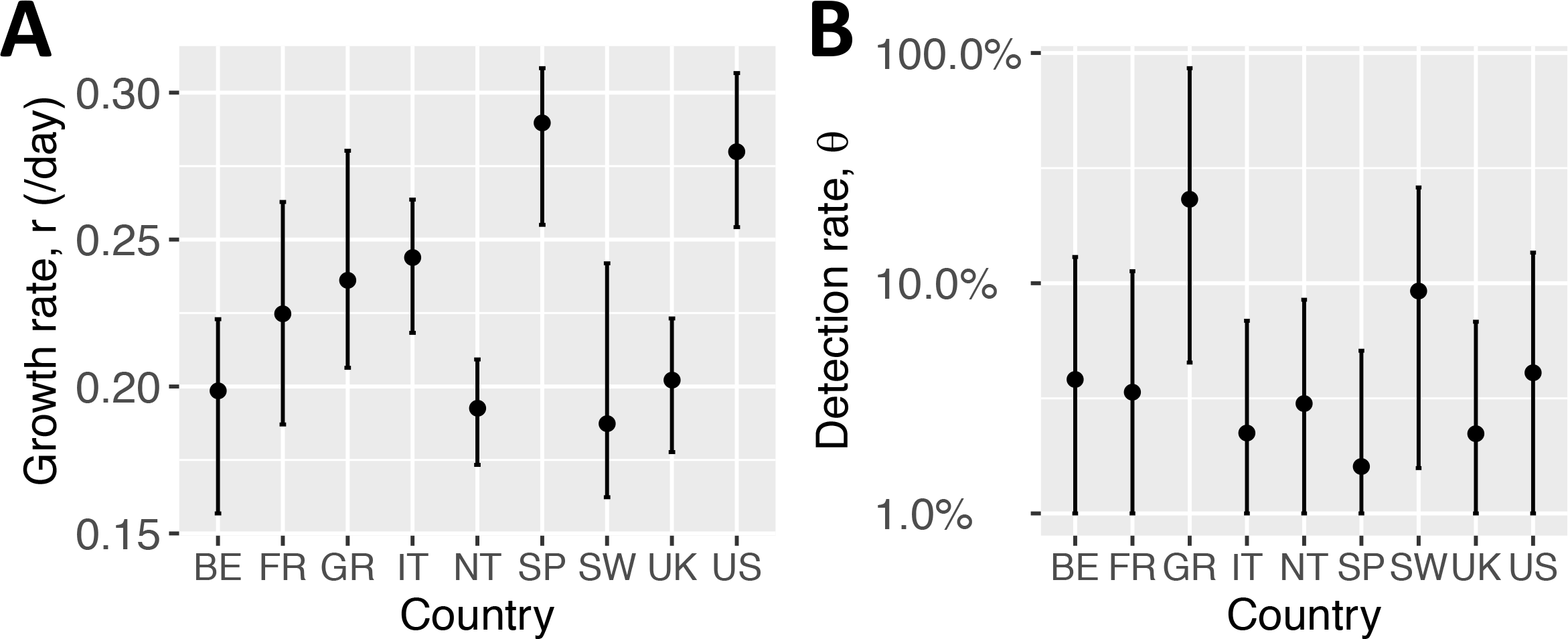
Point estimates and estimated ranges of the exponential growth rate, *r* and the detection rate, *θ*, in each country. BE: Belgium; FR: France; GR: Germany; IT: Italy; NT: Netherlands; SP: Spain; SW: Switzerland; UK: United Kingdom; US: United States.

We next estimated the detection rate, i.e. the probability of an infected person being identified by surveillance. Assuming a constant detection rate during the period when data used for inference are collected, we found that Germany, Switzerland and the US have point estimates of the detection rate at 58%, 20% and 12%, respectively. In the other countries examined, the detection rates were very low (less than 10%). This provides a natural explanation of the high number of reported cases compared to the relative low number of deaths in Germany during March 2020.

We caution that unlike the epidemic growth rate which is constrained by the time series of death count, the estimation of detection rate is highly dependent on the fixed parameter values assumed in the model, such as the infection fatality ratio, and the time from infection to case confirmation. If the infection fatality ratio is lower than we assumed, we would estimate even lower detection rates. To fully assess the uncertainties in the estimation, we performed sensitivity analysis varying parameter values within ranges based on the best current knowledge (see Supplementary Material). In general, the dection rate cannot be quantified precisely; however, the detection rates are likely less than 25% except for Germany and Switzerland (Fig. 2). Although large uncertainties exist, we find that the detection rate in Germany is much higher than the rates in the other countries. Overall, our results show that even in countries with well-developed medical and public health infrastructures, the detection rate for COVID-19 is in general low, likely due to the high percentage of infected individuals with no or mild-to-moderate symptoms(*9, 10*). This emphasize the importance of wearing personal protective equipment to prevent potential transmission from the large population of unidentified individuals, and more aggressive testing and contact tracing are needed to identify most infected individuals.

We further tested the possibility of changes in surveillance intensity, and found no statistical support (Table S2). We emphasize that this conclusion only applies to the period when the incidence data used for estimation were collected (Table S1). It is likely that the surveillance intensity was different during other periods of the outbreaks. As shown in Fig. 1, in all countries, the red open circles, i.e. data that were not used for inference, are mostly below the red band predicted by our model during very early outbreak. This indicates that in these countries, the detection rates were even lower than we estimated such that there were very few cases detected, although thousands of infected individuals were already infected. Some of the red open circles in the US seems during late March are above the red band, suggesting increases in surveillance intensity after the time period of our data collection.

### Implications to intervention strategies - ‘hit hard, hit early’

Using our empirical estimates of the growth rates, we explored the implications for public health efforts needed to control the COVID-19 outbreak. We considered an outbreak scenario in a city with a population size of 10 million. Because it may take at least one and a half years for an effective vaccine to be developed and deployed, below we compared outbreak outcomes under different scenarios at 18 months.

We first calculated the length of time for the epidemic to reach epidemic peak, assuming only one infected individual at day 0. When the growth rate is higher than 0.027/day, the epidemic peak will occur in less than 18 months and a large fraction of the population (>50%) will be infected. If our goal is that the total fraction of infected individuals is less than 10% at the end of 18 months, the growth rate has to be less than 0.025/day (i.e. an extremely slow growth rate with a doubling time of more than 28 days; Fig. 3B). This suggests that moderate social distancing efforts will be insufficient to delay the epidemic peak beyond 18 months. On the other hand, if these targeted growth rates are achieved through very strong public health interventions, a little more effort would lead to enormous public health benefit, i.e. the total infected fraction decreases exponentially when *r* decrease beyond 0.023/day as shown in Fig. 3B.

**Figure 3.**
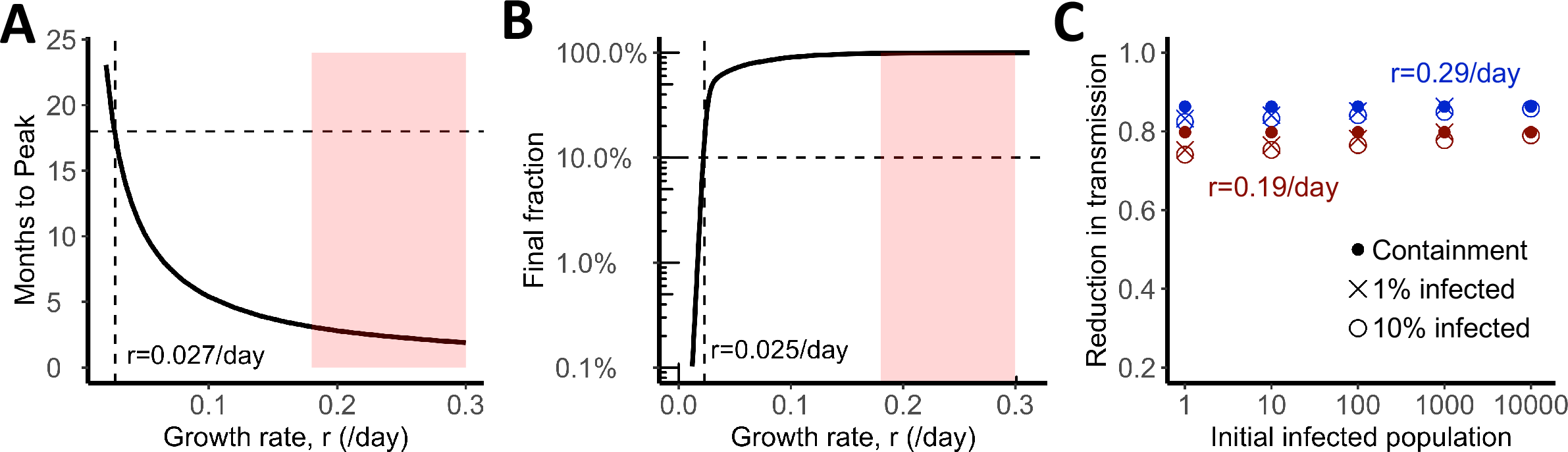
Strong control measures are needed to achieve measurable benefits, no matter the goal is mitigation (‘flatten the curve’) or containment. **(A)** Predicted time to epidemic peak for different growth rates. To delay epidemic peak beyond 18 months, a growth rate less than 0.027/day is needed. This threshold is denoted by dashed lines. Red area shows the ranges of growth rates estimated for the eight European countries, the US and Wuhan, China. **(B)** Final fraction of infected individuals after 18 months. A growth rate less than 0.022/day (doubling time more than 32 days) is needed to achieve the goal that less than 10% of individuals are infected (dashed lines). However, the benefit, i.e. fraction of uninfected individuals, increases exponentially when the growth rate is further reduced beyond the threshold. **(C)** Similar levels of efforts, measured as fractions of transmission reduction, are needed to achieve containment, i.e. reverting epidemic growth (dots), or mitigation, i.e. the final fraction of infected individuals is 1% (x) or 10% (open circle). We assumed epidemic growth rates of 0.19 (red) or 0.28/day (blue).

To further corroborates our results, we calculated the efforts needed to achieve three goals in 18 months: 1) virus containment, i.e. epidemic stops growing, 2) the total infected population is 10%, and 3) the total infected population is 1%. We found that the efforts needed are similar, especially when the population of infected individuals is already more than 100 (Fig. 3C), i.e. a scenario that many cities around the globe are facing now. For example, when an outbreak grows at rate 0.28/day (as we estimated for the US), the levels of efforts needed to achieve the three goals are between 84% and 86% reduction in transmission; whereas when the growth rate is 0.19/day, the levels of effort needed are between 74% and 80% reduction. These high levels of reduction needed argue for very strong and comprehensive intervention efforts implemented as soon as possible, no matter whether the goal is containment or mitigation - a strategy reminiscent of the ‘hit hard, hit early’ paradigm in treating HIV infection in a patient (*15*).

## Discussion

The epidemic growth rate for disease spread depends on many factors, including biological(*16*), demographic, and social factors. In this work, we report high COVID-19 epidemic growth rates between 0.19-0.29/day and short doubling times between 2.4-3.7 days across the eight most affected countries in Europe and in the US (as of March 31, 2020). This is consistent with our previous estimate for the early COVID-19 outbreak in Wuhan, China(*6*). Altogether, these results demonstrate COVID-19 to be a highly transmissible disease in the absence of strong control measurements irrespective of heterogeneities in geographic and social settings. We also find that most of infected individuals are not identified/detected, similar to findings in Wuhan, China(*6, 11*). This has important implications to both pharmaceutical and non-pharmaceutical interventions as we discuss below.

First, with the global efforts to develop vaccines for COVID-19, it is important to have an accurate measure of the basic reproductive number, *R*_0_, to set the threshold level of herd immunity needed to prevent transmission, i.e. 1-1/*R*_0_. Previously, *R*_0_ for COVID-19 were estimated to be between 2-3 (*2-5*) and were widely reported in official documents (*17*) and public media. These estimates are mostly based on an epidemic growth rate between 0.1 and 0.14/day (*2-5*), which is inconsistent with many new lines of evidence and data as discussed in this paper and a previous work (*6*). Previously we showed that with a mean serial interval, defined as the duration of time between onset of symptoms in an index case and a secondary case, of 6-9 days (*3, 18*) and an epidemic growth rate between 0.19-0.29/day, the value of *R*_0_ must be higher than 3 (*6*). Using the same framework as in Ref. 6, we find that when the growth rate is 0.19/day, the median *R*_0_ is estimated to be 4.0 (95% CI: 3.1 and 5.0); whereas when the growth rate is 0.29/day, the median *R*_0_ is estimated to be 7.1 (95% CI: 5.1 and 9.6). Although shorter serial intervals are reported in the literature (*19, 20*), it was noted that this is likely due to strong intervention efforts(*18, 19*). Given the potentially long infectious period in individuals with mild or severe symptoms (*21*), the mean serial interval during early outbreak in the absence of strong intervention (a likely scenario in most countries examined here) is unlikely less than 6 days. Overall, our results imply that a large fraction of the population needs to be vaccinated if an effective vaccine is to prevent the spread of the virus. In addition, if the virus is allowed to spread through the population, a large fraction of the population (e.g. 75% and 86% for an *R*_0_ of 4.0 and 7.1, repectively) will be infected even if the epidemic growth curve is flattened by control efforts.

Second, the awareness of the extraordinary high rates of COVID-19 spread during the current outbreak is critically important for epidemic preparedness. This is because the short doubling time means that health care systems can be overwhelmed in a couple of weeks rather than several months in the absence of control. A recent report shows that the number of COVID-19 patients admitted to intensive care units (ICUs) in Italy grew at a rate of approximately 0.25/day(*22*), remarkably consistent with our estimates. With such a high growth rate, there was only a very short window period for preparation (*22*). Of course, heterogeneities in the growth rate may exist among different areas within each a country. We note that our inference is largely driven by data collected from highly populated areas, such as Wuhan in China, Lombardy in Italy, and New York city in the US. Further work is needed to assess heterogeneity in the rate of spread across areas with different population densities.

Third, the high epidemic growth rates suggest that moderate control efforts will not sufficiently slow the virus spread to achieve measurable public health benefits. This may explain the continuous growth of the outbreak in some countries despite measures, such as work and school closures, were in place. We found that similarly high levels of efforts to reduce overall transmission by 74-86% are needed to delay the epidemic peak and protect a large fraction of the population from infection in 18 months (mitigation) or to reduce *R*_0_ below 1 (containment).

Lastly, our finding that the majority of infected individuals are not identified suggests that in most infected individuals, the symptoms are likely to be mild, and many of them may not be aware of their infection status. This argues for extensive, universally available testing to identify and isolate most infected individuals as well as the use of personal protective equipment to prevent potential transmission from individuals with no or mild symptoms.

Overall, in the absence of very strong control, the virus can cause high mortality and morbidity (*23, 24*) due to the high number of expected infections, which places an extremely heavy burden on even the most advanced health care systems (*24, 25*). Thus, with COVID-19, half-measures will not be effective in meeting public health goals. To delay the peak or to reverse the growth of the epidemic, we probably need all feasible tools available, i.e. extensive testing, isolation and quarantine, use of personal protective equipment, coupled with comprehensive social distancing. This is a strategy reminiscent of the ‘hit hard, hit early’ paradigm in treating HIV infected individuals (*15*). China, South Korea, and Singapore have proven that containment is possible with appropriate measures. Because there will be extensive economic impacts of the global COVID-19 pandemic regardless of how principled our response is, the question is not balancing public health with damage to the economy, but rather how many lives can we save.

## Data Availability

All data is available in the main text or the supplementary materials.

## Acknowledgments

We would like to thank Alan Perelson for suggestions and critical reading of the manuscript. The work is partially funded by the Laboratory Directed Research and Development (LDRD) Rapid Response Program through the Center for Nonlinear Studies at Los Alamos National Laboratory. RK and SS would like to acknowledge funding from DARPA (HR0011938513), the Center for Nonlinear Studies. ERS was funded though NIH grant (R01AI135946).

## Author contributions

RK and NH conceived the project; RK performed literature search and designed the study; SS collected data; RK, SS and ERS performed analyses; RK, SS, ERS and NH wrote and edited the manuscript.

